# A rapid systematic review of the effectiveness of out-of-hours palliative care telephone advice lines for people living at home and their carers

**DOI:** 10.1101/2023.08.30.23294814

**Authors:** Therese Johansson, Rachel L. Chambers, Thomas Curtis, Sophie Pask, Sarah Greenley, Molly Brittain, Anna E. Bone, Lynn Laidlaw, Ikumi Okamoto, Stephen Barclay, Irene Higginson, Fliss E. M. Murtagh, Katherine E. Sleeman

## Abstract

**Background:** People with palliative and end-of-life care needs in the community and their carers often rely on out-of-hours services to remain at home. Policymakers internationally have recommended implementation of telephone advice lines to ensure 24-hour access to support. However, little is known about their effectiveness.

**Aim:** To review the evidence for the clinical and cost effectiveness of out-of-hours-telephone advice lines for adults with palliative care needs living at home and their carers, and report service characteristics associated with effectiveness.

**Design:** Rapid systematic review, with narrative synthesis (PROSPERO ID: CRD42023400370).

**Data sources:** Three databases (Medline, EMBASE, CINAHL) were searched in February 2023 for studies reporting on telephone advice lines with at least partial out-of-hours availability. Study quality was assessed using the Mixed Methods Appraisal Tool.

**Results:** Twenty-one studies, published 2000-2022, were included. Most studies were observational; none were experimental. Three were comparative, and seven lacked explicit research questions or methods. Results were largely descriptive, focusing on service development and use, and process measures. Patient and carer outcomes were primarily reported qualitatively. Only two studies investigated possible system outcomes, by examining care-seeking behaviour after using telephone advice lines.

**Conclusion:** Existing evidence for the effectiveness of telephone advice lines is limited. The lack of experimental studies evaluating individual or system-level outcomes prevents assessment of the effectiveness/cost-effectiveness of service models. There is a clear need for more rigorous evaluations using consistent reporting, and inclusion of patient and carer perspectives during both development and implementation. Recommendations for future evaluations are provided.

**Key statements:** *What is already known about the topic?:*

- Urgent and unplanned emergency department and hospital admission is frequent for people in the final months of life.
- Designated palliative care telephone advice lines have been recommended internationally to ensure round-the-clock access to support from trained professionals and are proposed to help reduce urgent and unplanned use of acute services.
- While a range of palliative care telephone advice lines exist, the evidence base for their effectiveness, in terms of patient and service use outcomes, is not known.

*What this paper adds:*

- This review provides an overview of published articles reporting palliative care telephone advice line models that have been developed and implemented.
- We demonstrate that existing research evidence for the effectiveness of telephone advice lines is limited and largely based on observational studies of insufficient methodological quality.
- Our synthesis of findings suggests telephone advice lines can offer guidance and reassurance that supports family carers in providing care at home for patients who prefer to die at home.

*Implications for practice, theory or policy:*

- Future development and evaluation of telephone advice lines need to include patients, carers, and other stakeholders to better understand what needs and preferences should shape the services.
- To address the limited and variable evidence identified, we provide recommendations for key components of structure and use of telephone advice line models that should be included in future research.

## Background

People with palliative and end-of-life care needs living at home and their carers often rely on out-of-hours service providers^1, 2^, particularly during periods of worsening symptoms, or changes in the individual’s condition^3^.

Unscheduled care and out-of-hours care services are frequently accessed by people approaching the end of life^1, 4, 5^, with emergency care utilisation rising considerably in the final month of life^6^. Still, a large proportion of palliative care is delivered in the community^7, 8^. Among those who die, almost 30% are seen by out-of-hours primary care services during their last 30 days of life^9^.

Timely access to palliative care support and advice from trained professionals is essential to support patients who prefer to be cared for, and die, at home (and their informal carers) who might face a complex array of symptoms and care needs^10, 11^. Despite international trends of increasing home deaths and the rising need for community-based palliative care^12^, access and provision of palliative care in the community is highly geographically variable^8, 13, 14^.

Palliative care telephone advice lines have been proposed internationally to provide timely support for people with palliative care needs and their carers, especially in rural areas^15–17^. In 2004, the UK National Institute for Health and Care Excellence has recommended provision of designated telephone advice lines to ensure that patients with life-limiting illnesses and their carers have round-the-clock access to support from trained palliative care professionals^18^. Almost 20 years later, however, designated palliative care telephone advice lines are not available everywhere in the UK, with 27% of areas having no access and 42% having only partial access during out-of-hours periods^19^. Consequently, many people rely on the national emergency numbers (i.e., 999 or NHS 111) and primary care service provision from out-of-hours General Practitioners and Community Nurses during these times. Though a wide variety of palliative care telephone advice lines are in use and development^19^, the effectiveness and cost-effectiveness of different service models has not been synthesised and the evidence base for their effectiveness and optimal implementation is limited.

### Aim

Our aim was to review the evidence for the clinical and cost-effectiveness of out-of-hours telephone advice lines for adults with palliative care needs living at home and their carers, and report how service characteristics are associated with effectiveness.

## Methods

### Study design

This rapid systematic review was planned and conducted in line with guidelines set out by Cochrane Rapid Reviews Methods Group^20^ and the World Health Organization^21^. The rapid review procedure is methodologically similar to a full systematic review but limits the scope and analytic process to provide a timely appraisal of existing evidence, e.g., by assigning a single reviewer for screening or limiting sources beyond electronic databases^22^. The reporting adheres to the Preferred Reporting Items for Systematic Reviews and Meta-Analyses (PRISMA) 2020 checklist^23^. A protocol was developed prior to commencing the study and made publicly available on PROSPERO (CRD42023400370).

### Eligibility criteria

Any published study reporting on the use, effect, or implementation of a telephone advice line available specifically for home-based adults with palliative care needs and their carers during at least part of the out-of-hours period was eligible, even if it was not the primary focus of an intervention. A broad definition was used to allow assessment of various types of telephone advice lines, provided there was a ‘reactive’ component (i.e., contact is patient/carer-initiated, as opposed to pre-booked appointments). Detailed inclusion and exclusion criteria are shown in Table 1.

**Table 1.** Inclusion/exclusion criteria.

|  | *Inclusion criteria* | *Exclusion criteria* |
| --- | --- | --- |
| *Population* | Study populations comprising adults (i.e., aged $\geq 18$) living at home with palliative care needs and family/unpaid carers | Study populations solely comprising:<br>- children or adolescents (i.e., aged $\leq 18$)<br>- adult carers of paediatric patients<br>- people without palliative care needs |
| *Exposure* | Any telephone advice line that is:<br>- available through telephone calls<br>- designated for providing palliative care support, help, and information<br>- reactive, i.e., contact is patient/carer-initiated<br>- accessible during at least parts of the out-of-hours periods (weekdays between 18.30-08.00, weekends, and public holidays) | Any telephone advice line that is:<br>- solely for health and social care professionals<br>- designated for patients in institutional care settings<br>- only available in-hours (weekdays between 08.00-18.30)<br>- only providing bereavement support |
| *Outcomes* | - All outcomes related to patients, carers, services, and costs, provided they are directly related to the telephone advice line | - No study will be excluded based on reported outcomes |
| *Study type* | - Peer-reviewed publications of all types (with or without a comparator) reporting data related to implementation or use of telephone advice lines | - Studies reporting only review-level evidence<br>- Study protocols<br>- Opinion pieces or editorials<br>- Conference abstracts |

### Search strategy

The searches were performed with assistance of an information specialist (SG) at Hull York Medical School, University of Hull. The search was developed in Medline by examining the indexing of known potentially relevant studies, designing, and testing a search using these studies before translation into other databases. Three databases were searched: Medline All from 1946, EMBASE from 1974 (via OVID), and CINAHL Complete from 1937 via EbscoHost. The search used for Medline incorporated elements from previous strategies for palliative care by Rietjens et al. ^24^ and Sladek et al^25^, combining concepts of palliative care and out-of-hours telephone advice lines using both database specific indexed terms and free text terms. Filters were used to only retrieve primary studies written in English and to remove conference abstracts, books, and editorials. No date limits were applied. All searches were performed on February 9, 2023. The full search strategies for all databases are provided in supplementary file 1. Database results were downloaded into EndNote and duplicates were removed using a recognised method^26^ before being uploaded into Covidence^27^.

### Study selection

The first author (TJ) screened titles and abstracts against the inclusion criteria using Covidence. To ensure screening consistency, a subsample was screened by a second reviewer (RC) and any conflicts were resolved by discussion. Full-text screening was performed independently by two reviewers (TJ and RC/MB), with any disagreements resolved by a third reviewer (AB).

### Data extraction

A template for data extraction was constructed. Data were extracted by one reviewer (TJ) and checked by a second (RC/MB/TC). Extracted data were collated using Microsoft Excel. Data for clinical and cost effectiveness included any effects of the telephone advice lines related to impact on patients’ and carers’ health and wellbeing, symptom management, coping, caring capacity, emergency service use and hospitalisation, place of care, place of death, and experiences and satisfaction of service use. Data for cost effectiveness included: costs of providing the telephone advice line and related resource use (i.e., costs related to care service use). Due to the heterogeneity of study designs and reported outcomes, studies were not excluded from synthesis due to missing data.

### Data synthesis

The quality of included studies was assessed using the *Mixed Methods Appraisal Tool* ^28^, which yields a score ranging 1-5 that represents the number of quality criteria met for the specific study type. Given the scarcity of literature, all included studies were considered relevant for the review aims and were retained for synthesis regardless of their quality appraisal score.

Due to heterogeneity in aims, methods, and outcomes of included studies, study findings were synthesized narratively^29^. The synthesis was guided by Donabedian’s three-component framework for evaluating quality of care^30^, which encompasses *structures* (physical resources and staff to ensure the intervention is delivered), *processes* (how structural components are used and transactions between care recipients and providers), and *outcomes* (effects on patients’ and carers’ health and wellbeing, health-related behaviours, and satisfaction with care).

Findings relating to telephone advice line characteristics, usage, and reported study outcomes were mapped onto Donabedian’s framework^30^. Since this review presents outcomes of clinical effectiveness at a population level, we differentiated outcomes of care as *patient/carer outcomes* (i.e., health status, wellbeing, and coping), and *health care system outcomes* (i.e., impact on the use of care services). Although Donabedian’s framework conceptualizes patient satisfaction and experience as partly separate to outcomes in care quality assessment, it was incorporated here as a facet of patient/carer outcomes as it is influenced by patient and environmental factors^31^.

Findings from the review were discussed with the project patient and public involvement (PPI) advisory group, as well as in an online workshop with ten volunteers identified through the PPI forums of King’s College London and University of Hull to help shape the synthesis^32^. Advice from PPI members was sought on interpreting key messages, understanding essential characteristics of advice lines, and how the findings resonated with their lived experience.

## Results

The database searches identified 622 articles. After removing duplicates, 388 articles remained for screening based on title and abstract (of which 80 randomly selected articles were dual screened). In total, 340 articles were excluded, leaving 48 for full text review. Twenty-one articles were included in the review. The PRISMA flow chart is presented in Figure 1, stating reasons for exclusion at each stage.

**Figure 1.**
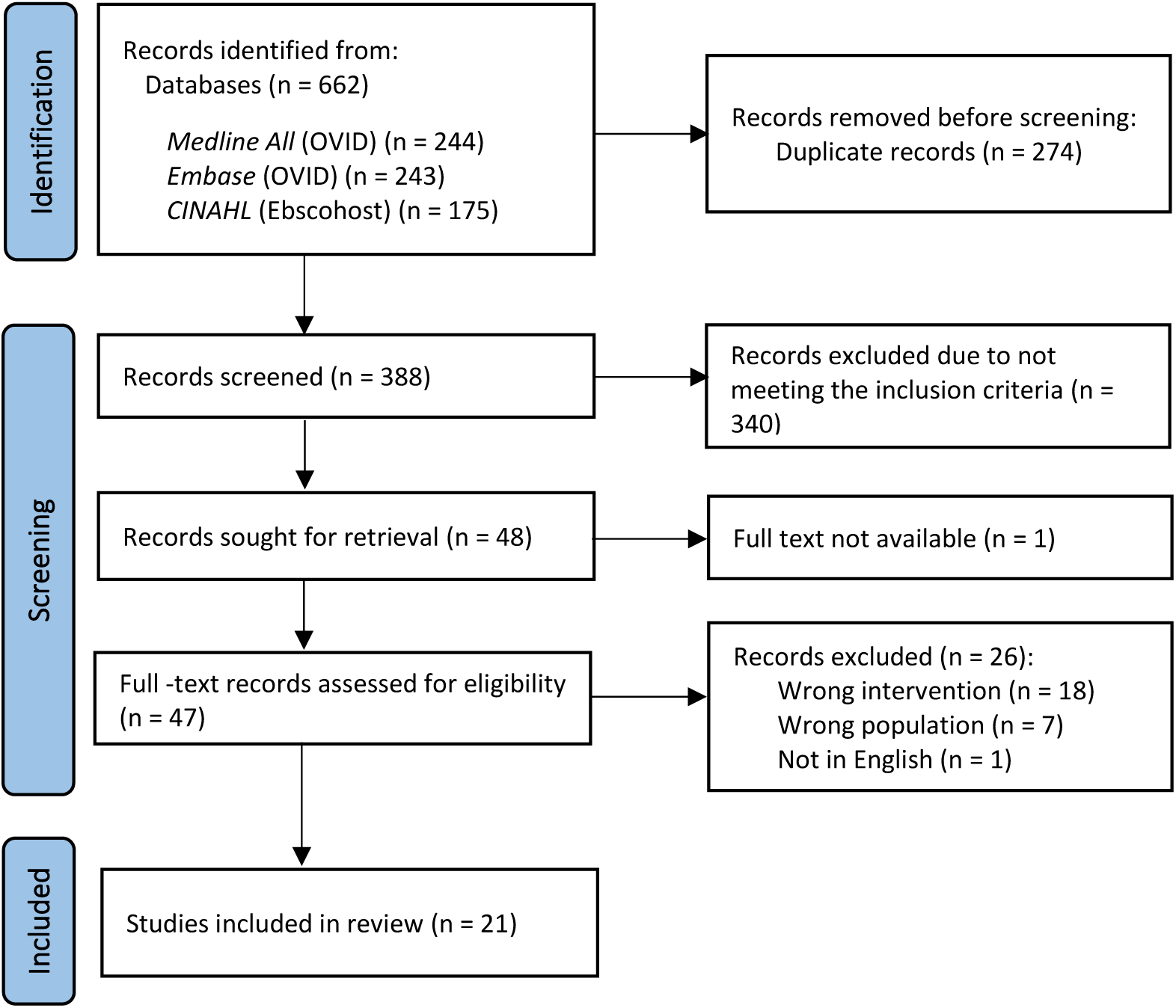
PRISMA flow diagram for systematic literature search results.

### Study descriptives

The 21 included studies were published between 2000 and 2022. Around half were conducted in the United Kingdom (*n*=11) ^33–43^, with the remaining studies coming from Australia (*n*=4) ^44–47^, the United States (*n*=2) ^48, 49^, Canada (*n*=1) ^50^, The Netherlands (*n*=1) ^51^, Singapore (*n*=1) ^52^ and Taiwan (*n*=1) ^53^.

Eight of the studies were quantitative (of which three were comparative) ^41, 44, 45, 48, 49, 51–53^, five used mixed-methods^35, 36, 38, 39, 47^, and one was qualitative^42^. Seven studies did not specify a methodology. Of these, four reported quantitative and qualitative data^34, 37, 40, 50^, and three presented quantitative data only^33, 43, 46^.

Most studies were retrospective observational studies or audits, using quantitative data either from existing call logs or purpose-built proformas for incoming calls to the telephone advice line as the primary source of data (*n*=15). Qualitative data were usually collected through feedback surveys or follow-up interviews with a sub-sample of callers. Table 2 presents the methodology of included studies.

**Table 2.**
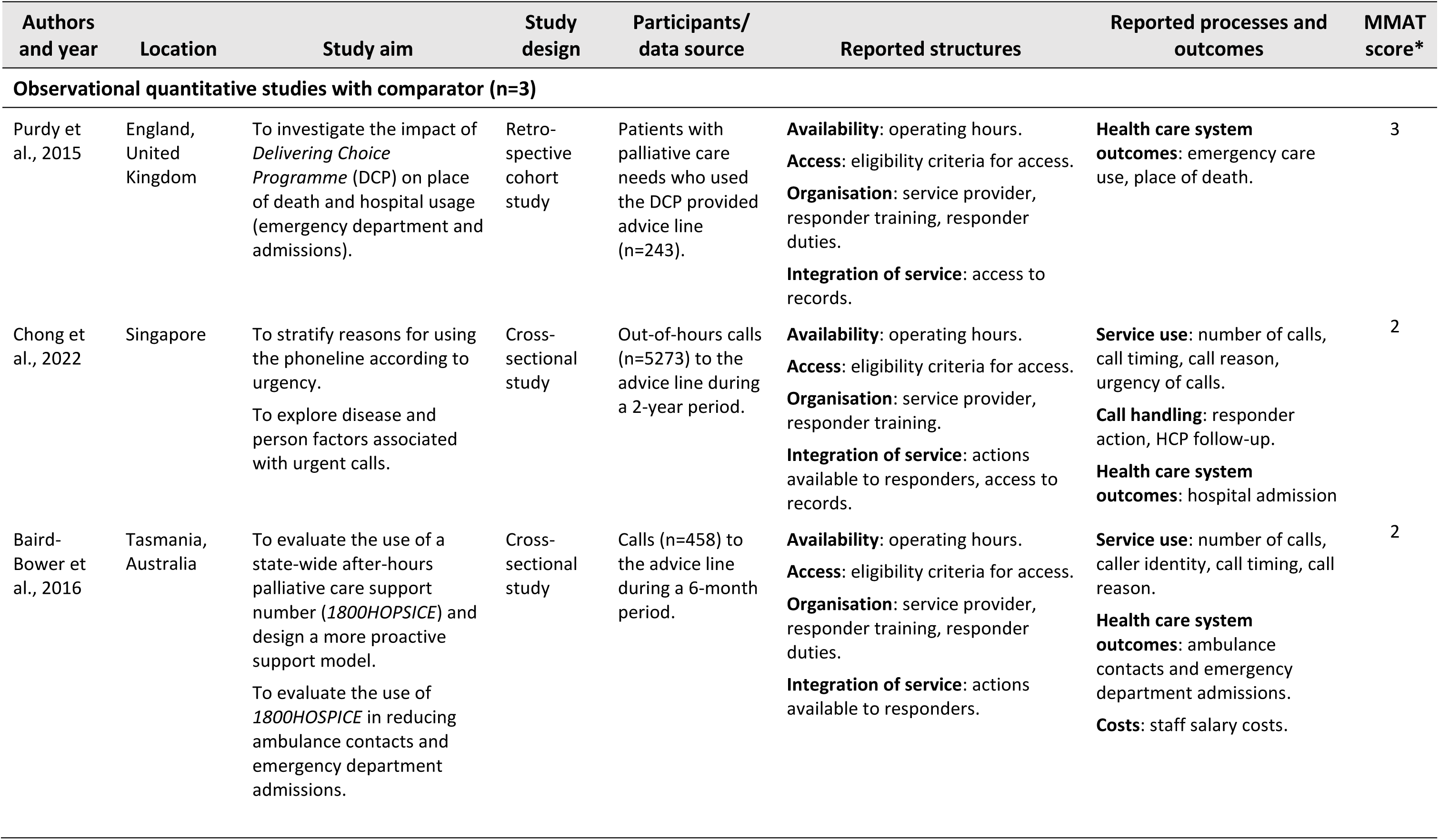
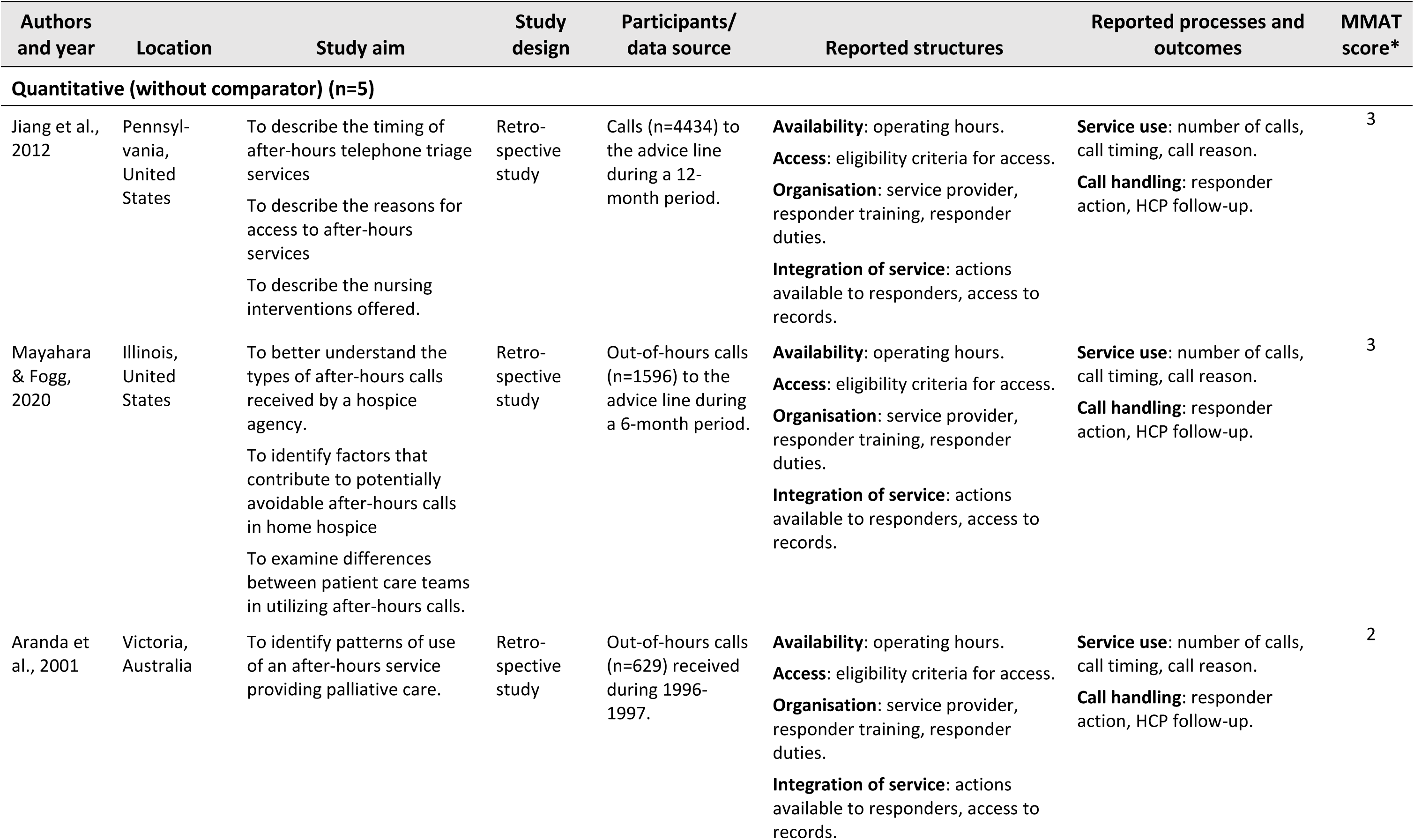
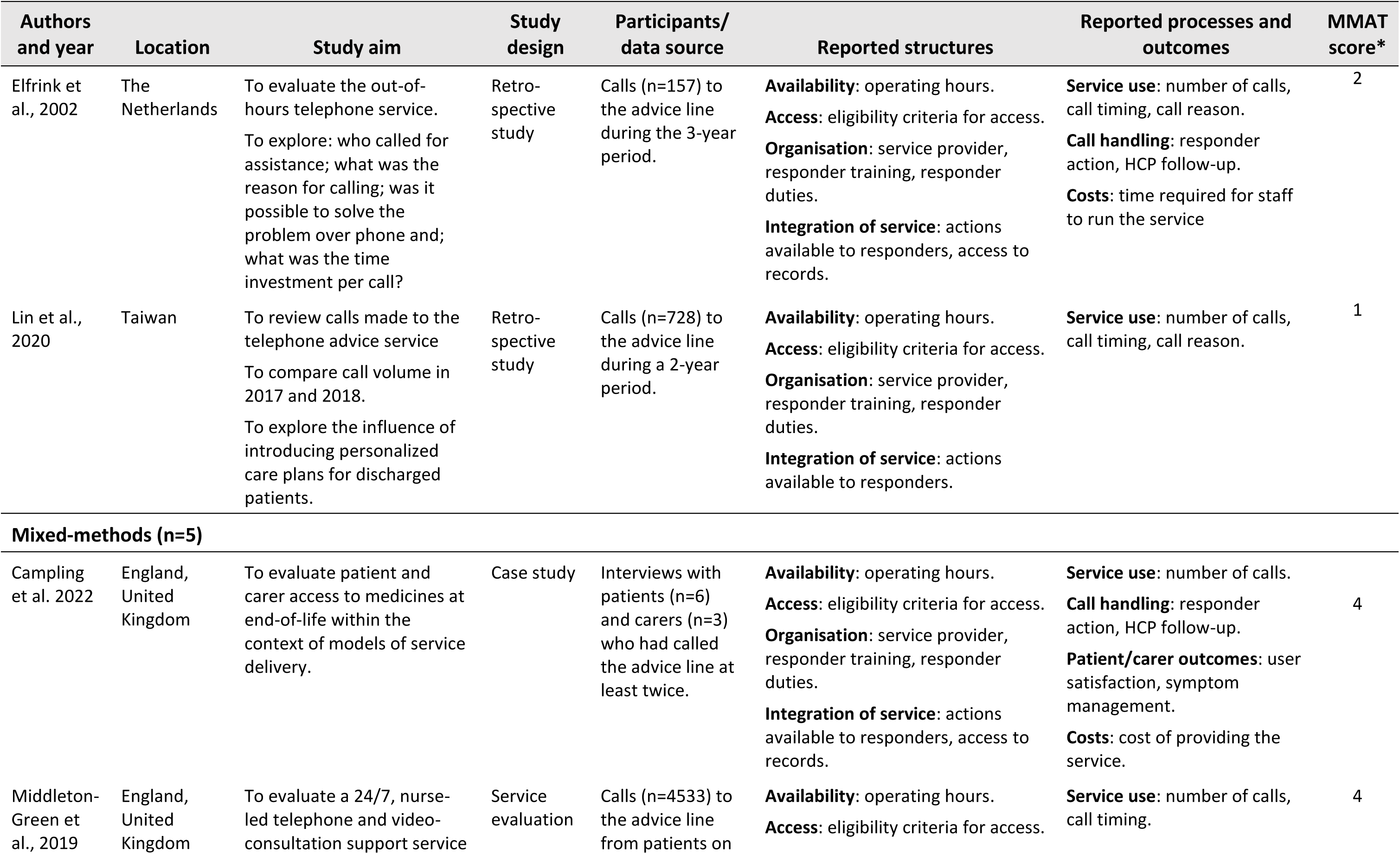
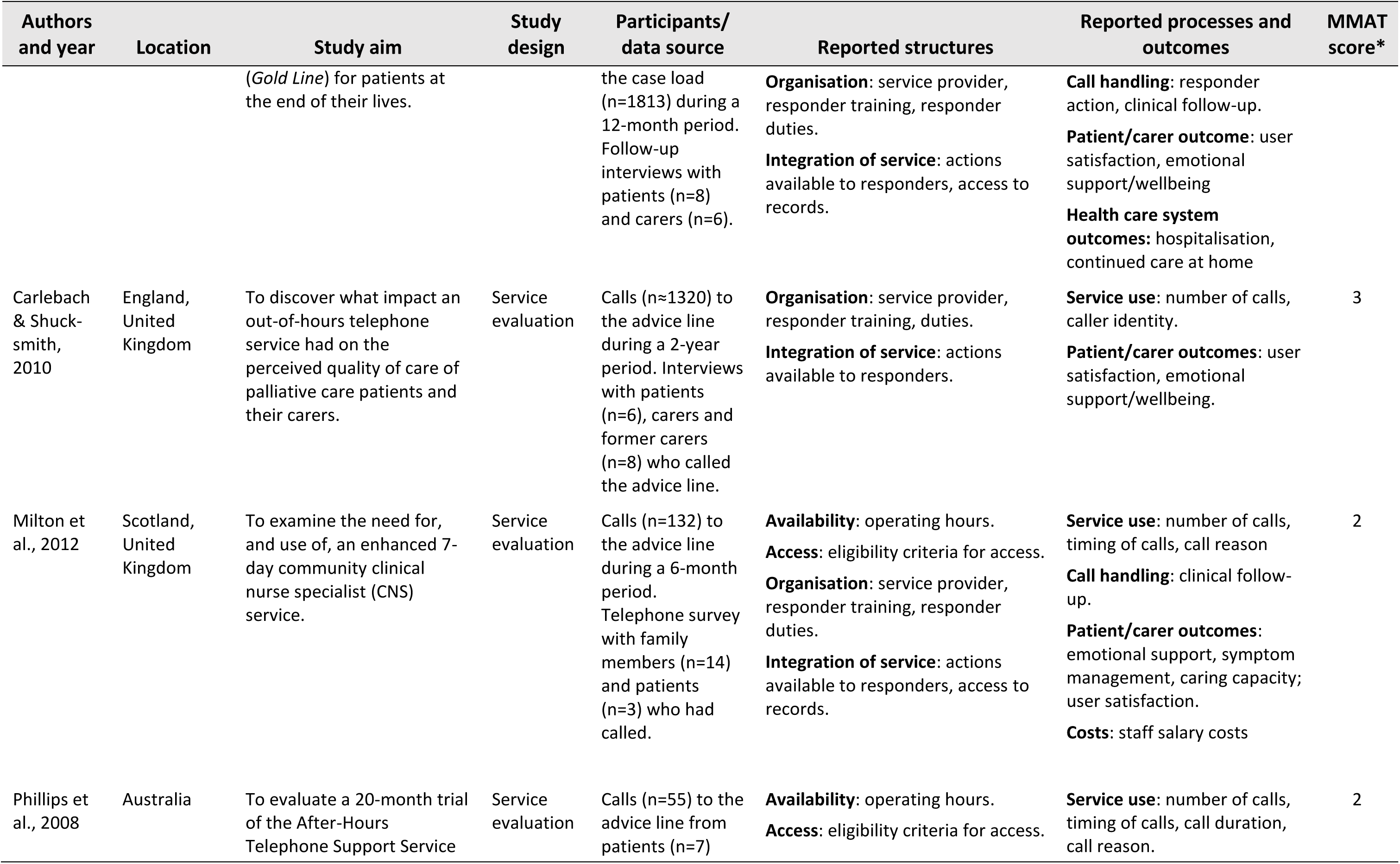
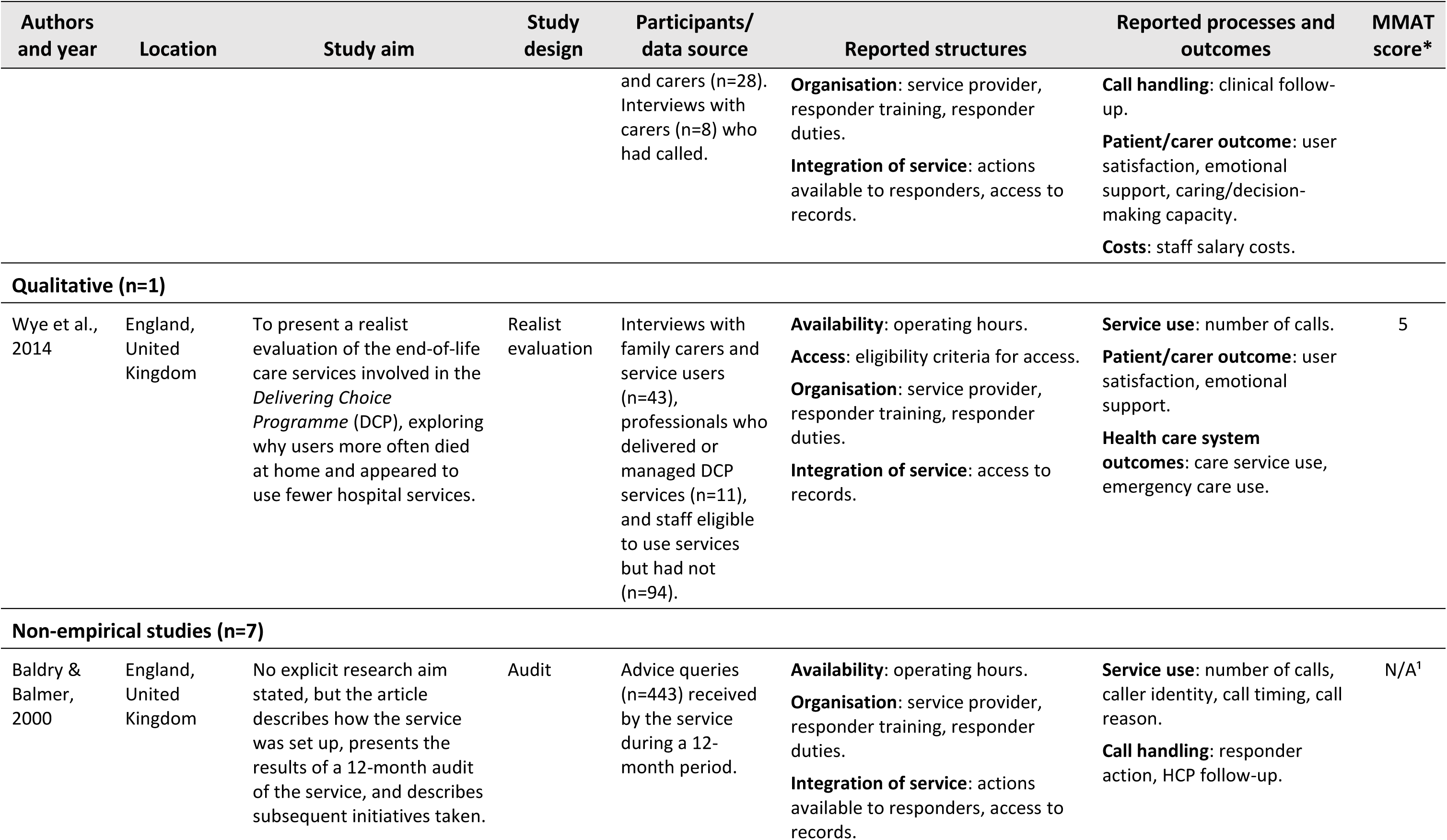
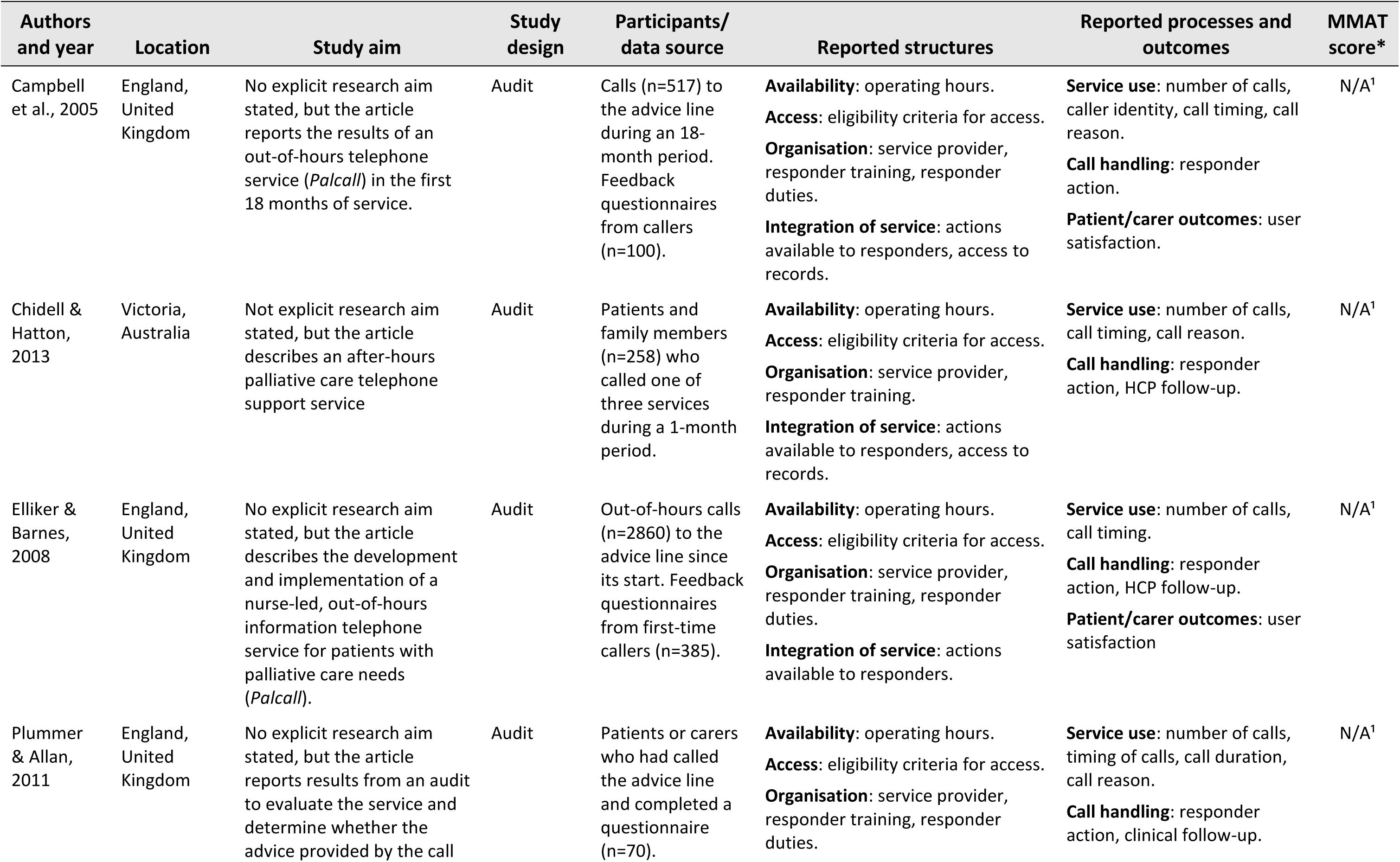
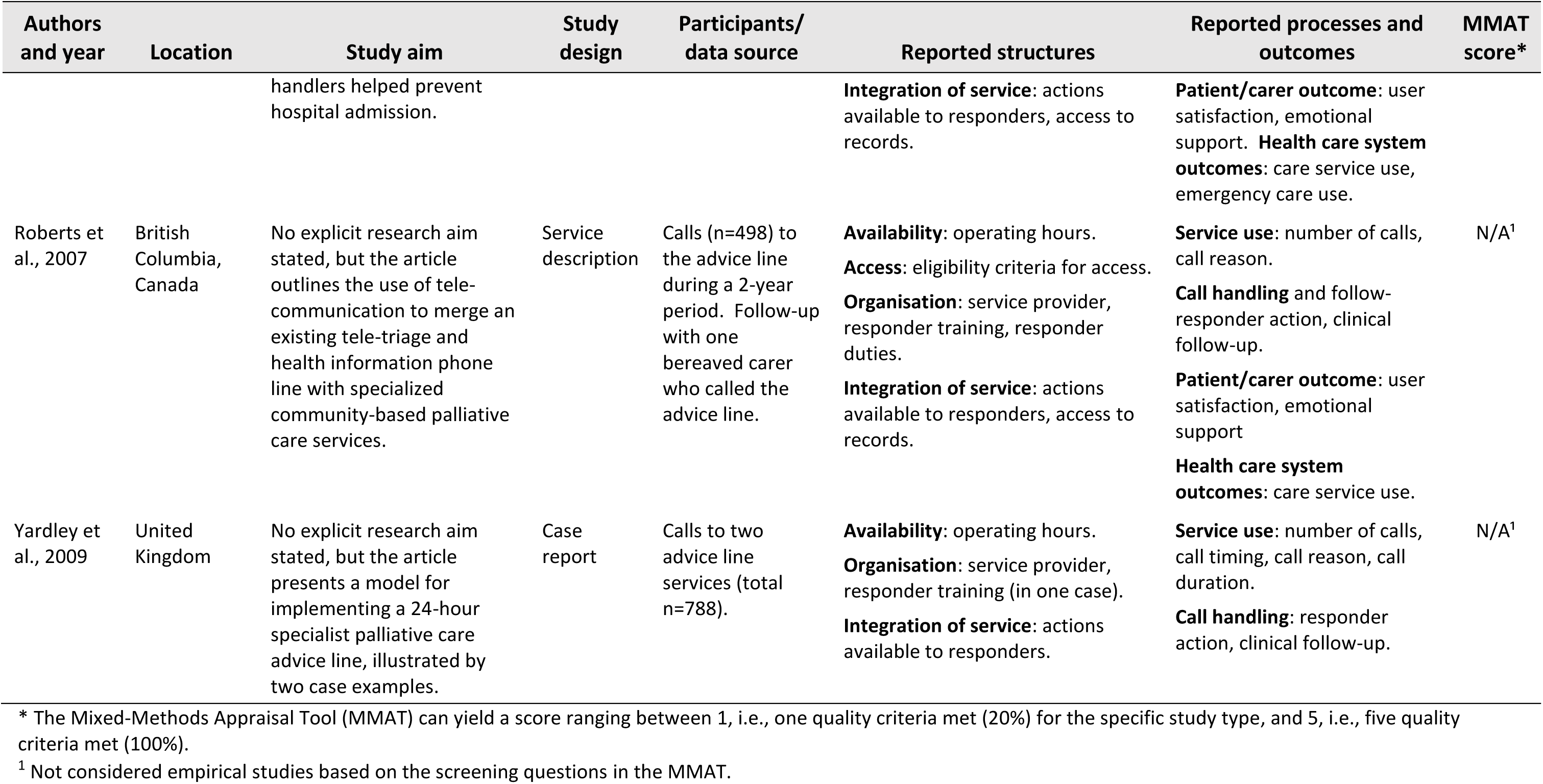
Overview of the methodology of included studies, grouped according to study type and ordered according to quality appraisal score.

| Authors and year | Location | Study aim | Study design | Participants/ data source | Reported structures | Reported processes and outcomes | MMAT score* |
| --- | --- | --- | --- | --- | --- | --- | --- |
| **Observational quantitative studies with comparator (n=3)** |  |  |  |  |  |  |  |
| Purdy et al., 2015 | England, United Kingdom | To investigate the impact of *Delivering Choice Programme* (DCP) on place of death and hospital usage (emergency department and admissions). | Retro-spective cohort study | Patients with palliative care needs who used the DCP provided advice line (n=243). | **Availability:** operating hours.<br>**Access:** eligibility criteria for access.<br>**Organisation:** service provider, responder training, responder duties.<br>**Integration of service:** access to records. | **Health care system outcomes:** emergency care use, place of death. | 3 |
| Chong et al., 2022 | Singapore | To stratify reasons for using the phoneline according to urgency.<br>To explore disease and person factors associated with urgent calls. | Cross-sectional study | Out-of-hours calls (n=5273) to the advice line during a 2-year period. | **Availability:** operating hours.<br>**Access:** eligibility criteria for access.<br>**Organisation:** service provider, responder training.<br>**Integration of service:** actions available to responders, access to records. | **Service use:** number of calls, call timing, call reason, urgency of calls.<br>**Call handling:** responder action, HCP follow-up.<br>**Health care system outcomes:** hospital admission | 2 |
| Baird-Bower et al., 2016 | Tasmania, Australia | To evaluate the use of a state-wide after-hours palliative care support number (*1800HOPSICE*) and design a more proactive support model.<br>To evaluate the use of *1800HOSPICE* in reducing ambulance contacts and emergency department admissions. | Cross-sectional study | Calls (n=458) to the advice line during a 6-month period. | **Availability:** operating hours.<br>**Access:** eligibility criteria for access.<br>**Organisation:** service provider, responder training, responder duties.<br>**Integration of service:** actions available to responders. | **Service use:** number of calls, caller identity, call timing, call reason.<br>**Health care system outcomes:** ambulance contacts and emergency department admissions.<br>**Costs:** staff salary costs. | 2 |
| **Quantitative (without comparator) (n=5)** |  |  |  |  |  |  |  |
| Jiang et al., 2012 | Pennsylvania, United States | To describe the timing of after-hours telephone triage services<br>To describe the reasons for access to after-hours services<br>To describe the nursing interventions offered. | Retro-spective study | Calls (n=4434) to the advice line during a 12-month period. | **Availability:** operating hours.<br>**Access:** eligibility criteria for access.<br>**Organisation:** service provider, responder training, responder duties.<br>**Integration of service:** actions available to responders, access to records. | **Service use:** number of calls, call timing, call reason.<br>**Call handling:** responder action, HCP follow-up. | 3 |
| Mayahara & Fogg, 2020 | Illinois, United States | To better understand the types of after-hours calls received by a hospice agency.<br>To identify factors that contribute to potentially avoidable after-hours calls in home hospice<br>To examine differences between patient care teams in utilizing after-hours calls. | Retro-spective study | Out-of-hours calls (n=1596) to the advice line during a 6-month period. | **Availability:** operating hours.<br>**Access:** eligibility criteria for access.<br>**Organisation:** service provider, responder training, responder duties.<br>**Integration of service:** actions available to responders, access to records. | **Service use:** number of calls, call timing, call reason.<br>**Call handling:** responder action, HCP follow-up. | 3 |
| Aranda et al., 2001 | Victoria, Australia | To identify patterns of use of an after-hours service providing palliative care. | Retro-spective study | Out-of-hours calls (n=629) received during 1996-1997. | **Availability:** operating hours.<br>**Access:** eligibility criteria for access.<br>**Organisation:** service provider, responder training, responder duties.<br>**Integration of service:** actions available to responders, access to records. | **Service use:** number of calls, call timing, call reason.<br>**Call handling:** responder action, HCP follow-up. | 2 |
| Elfrink et al., 2002 | The Netherlands | To evaluate the out-of-hours telephone service.<br>To explore: who called for assistance; what was the reason for calling; was it possible to solve the problem over phone and; what was the time investment per call? | Retro-spective study | Calls (n=157) to the advice line during the 3-year period. | **Availability:** operating hours.<br>**Access:** eligibility criteria for access.<br>**Organisation:** service provider, responder training, responder duties.<br>**Integration of service:** actions available to responders, access to records. | **Service use:** number of calls, call timing, call reason.<br>**Call handling:** responder action, HCP follow-up.<br>**Costs:** time required for staff to run the service | 2 |
| Lin et al., 2020 | Taiwan | To review calls made to the telephone advice service<br>To compare call volume in 2017 and 2018.<br>To explore the influence of introducing personalized care plans for discharged patients. | Retro-spective study | Calls (n=728) to the advice line during a 2-year period. | **Availability:** operating hours.<br>**Access:** eligibility criteria for access.<br>**Organisation:** service provider, responder training, responder duties.<br>**Integration of service:** actions available to responders. | **Service use:** number of calls, call timing, call reason. | 1 |
| **Mixed-methods (n=5)** |  |  |  |  |  |  |  |
| Campling et al. 2022 | England, United Kingdom | To evaluate patient and carer access to medicines at end-of-life within the context of models of service delivery. | Case study | Interviews with patients (n=6) and carers (n=3) who had called the advice line at least twice. | **Availability:** operating hours.<br>**Access:** eligibility criteria for access.<br>**Organisation:** service provider, responder training, responder duties.<br>**Integration of service:** actions available to responders, access to records. | **Service use:** number of calls.<br>**Call handling:** responder action, HCP follow-up.<br>**Patient/carers outcomes:** user satisfaction, symptom management.<br>**Costs:** cost of providing the service. | 4 |
| Middleton-Green et al., 2019 | England, United Kingdom | To evaluate a 24/7, nurse-led telephone and video-consultation support service | Service evaluation | Calls (n=4533) to the advice line from patients on | **Availability:** operating hours.<br>**Access:** eligibility criteria for access. | **Service use:** number of calls, call timing. | 4 |

| Authors and year | Location | Study aim | Study design | Participants/<br>data source | Reported structures | Reported processes and outcomes | MMAT score* |
| --- | --- | --- | --- | --- | --- | --- | --- |
|  |  | (*Gold Line*) for patients at the end of their lives. |  | the case load (n=1813) during a 12-month period. Follow-up interviews with patients (n=8) and carers (n=6). | **Organisation:** service provider, responder training, responder duties.<br>**Integration of service:** actions available to responders, access to records. | **Call handling:** responder action, clinical follow-up.<br>**Patient/carers outcome:** user satisfaction, emotional support/wellbeing<br>**Health care system outcomes:** hospitalisation, continued care at home |  |
| Carlebach & Shuck-smith, 2010 | England, United Kingdom | To discover what impact an out-of-hours telephone service had on the perceived quality of care of palliative care patients and their carers. | Service evaluation | Calls (n≈1320) to the advice line during a 2-year period. Interviews with patients (n=6), carers and former carers (n=8) who called the advice line. | **Organisation:** service provider, responder training, duties.<br>**Integration of service:** actions available to responders. | **Service use:** number of calls, caller identity.<br>**Patient/carers outcomes:** user satisfaction, emotional support/wellbeing. | 3 |
| Milton et al., 2012 | Scotland, United Kingdom | To examine the need for, and use of, an enhanced 7-day community clinical nurse specialist (CNS) service. | Service evaluation | Calls (n=132) to the advice line during a 6-month period. Telephone survey with family members (n=14) and patients (n=3) who had called. | **Availability:** operating hours.<br>**Access:** eligibility criteria for access.<br>**Organisation:** service provider, responder training, responder duties.<br>**Integration of service:** actions available to responders, access to records. | **Service use:** number of calls, timing of calls, call reason<br>**Call handling:** clinical follow-up.<br>**Patient/carers outcomes:** emotional support, symptom management, caring capacity; user satisfaction.<br>**Costs:** staff salary costs | 2 |
| Phillips et al., 2008 | Australia | To evaluate a 20-month trial of the After-Hours Telephone Support Service | Service evaluation | Calls (n=55) to the advice line from patients (n=7) | **Availability:** operating hours.<br>**Access:** eligibility criteria for access. | **Service use:** number of calls, timing of calls, call duration, call reason. | 2 |
|  |  |  |  | and carers (n=28). Interviews with carers (n=8) who had called. | **Organisation:** service provider, responder training, responder duties.<br>**Integration of service:** actions available to responders, access to records. | **Call handling:** clinical follow-up.<br>**Patient/carers outcome:** user satisfaction, emotional support, caring/decision-making capacity.<br>**Costs:** staff salary costs. |  |
| **Qualitative (n=1)** |  |  |  |  |  |  |  |
| Wye et al., 2014 | England, United Kingdom | To present a realist evaluation of the end-of-life care services involved in the *Delivering Choice Programme* (DCP), exploring why users more often died at home and appeared to use fewer hospital services. | Realist evaluation | Interviews with family carers and service users (n=43), professionals who delivered or managed DCP services (n=11), and staff eligible to use services but had not (n=94). | **Availability:** operating hours.<br>**Access:** eligibility criteria for access.<br>**Organisation:** service provider, responder training, responder duties.<br>**Integration of service:** access to records. | **Service use:** number of calls.<br>**Patient/carers outcome:** user satisfaction, emotional support.<br>**Health care system outcomes:** care service use, emergency care use. | 5 |
| **Non-empirical studies (n=7)** |  |  |  |  |  |  |  |
| Baldry & Balmer, 2000 | England, United Kingdom | No explicit research aim stated, but the article describes how the service was set up, presents the results of a 12-month audit of the service, and describes subsequent initiatives taken. | Audit | Advice queries (n=443) received by the service during a 12-month period. | **Availability:** operating hours.<br>**Organisation:** service provider, responder training, responder duties.<br>**Integration of service:** actions available to responders, access to records. | **Service use:** number of calls, caller identity, call timing, call reason.<br>**Call handling:** responder action, HCP follow-up. | N/A<sup>1</sup> |
| Campbell et al., 2005 | England, United Kingdom | No explicit research aim stated, but the article reports the results of an out-of-hours telephone service (*Palcall*) in the first 18 months of service. | Audit | Calls (n=517) to the advice line during an 18-month period. Feedback questionnaires from callers (n=100). | **Availability:** operating hours.<br>**Access:** eligibility criteria for access.<br>**Organisation:** service provider, responder training, responder duties.<br>**Integration of service:** actions available to responders, access to records. | **Service use:** number of calls, caller identity, call timing, call reason.<br>**Call handling:** responder action.<br>**Patient/carer outcomes:** user satisfaction. | N/A<sup>1</sup> |
| Chidell & Hatton, 2013 | Victoria, Australia | Not explicit research aim stated, but the article describes an after-hours palliative care telephone support service | Audit | Patients and family members (n=258) who called one of three services during a 1-month period. | **Availability:** operating hours.<br>**Access:** eligibility criteria for access.<br>**Organisation:** service provider, responder training.<br>**Integration of service:** actions available to responders, access to records. | **Service use:** number of calls, call timing, call reason.<br>**Call handling:** responder action, HCP follow-up. | N/A<sup>1</sup> |
| Elliker & Barnes, 2008 | England, United Kingdom | No explicit research aim stated, but the article describes the development and implementation of a nurse-led, out-of-hours information telephone service for patients with palliative care needs (*Palcall*). | Audit | Out-of-hours calls (n=2860) to the advice line since its start. Feedback questionnaires from first-time callers (n=385). | **Availability:** operating hours.<br>**Access:** eligibility criteria for access.<br>**Organisation:** service provider, responder training, responder duties.<br>**Integration of service:** actions available to responders. | **Service use:** number of calls, call timing.<br>**Call handling:** responder action, HCP follow-up.<br>**Patient/carer outcomes:** user satisfaction | N/A<sup>1</sup> |
| Plummer & Allan, 2011 | England, United Kingdom | No explicit research aim stated, but the article reports results from an audit to evaluate the service and determine whether the advice provided by the call | Audit | Patients or carers who had called the advice line and completed a questionnaire (n=70). | **Availability:** operating hours.<br>**Access:** eligibility criteria for access.<br>**Organisation:** service provider, responder training, responder duties. | **Service use:** number of calls, timing of calls, call duration, call reason.<br>**Call handling:** responder action, clinical follow-up. | N/A<sup>1</sup> |
|  |  | handlers helped prevent hospital admission. |  |  | **Integration of service:** actions available to responders, access to records. | **Patient/carer outcome:** user satisfaction, emotional support. **Health care system outcomes:** care service use, emergency care use. |  |
| Roberts et al., 2007 | British Columbia, Canada | No explicit research aim stated, but the article outlines the use of tele-communication to merge an existing tele-triage and health information phone line with specialized community-based palliative care services. | Service description | Calls (n=498) to the advice line during a 2-year period. Follow-up with one bereaved carer who called the advice line. | **Availability:** operating hours.<br>**Access:** eligibility criteria for access.<br>**Organisation:** service provider, responder training, responder duties.<br>**Integration of service:** actions available to responders, access to records. | **Service use:** number of calls, call reason.<br>**Call handling** and follow-responder action, clinical follow-up.<br>**Patient/carer outcome:** user satisfaction, emotional support<br>**Health care system outcomes:** care service use. | N/A<sup>1</sup> |
| Yardley et al., 2009 | United Kingdom | No explicit research aim stated, but the article presents a model for implementing a 24-hour specialist palliative care advice line, illustrated by two case examples. | Case report | Calls to two advice line services (total n=788). | **Availability:** operating hours.<br>**Organisation:** service provider, responder training (in one case).<br>**Integration of service:** actions available to responders. | **Service use:** number of calls, call timing, call reason, call duration.<br>**Call handling:** responder action, clinical follow-up. | N/A<sup>1</sup> |

\* The Mixed-Methods Appraisal Tool (MMAT) can yield a score ranging between 1, i.e., one quality criteria met (20%) for the specific study type, and 5, i.e., five quality criteria met (100%).
<sup>1</sup> Not considered empirical studies based on the screening questions in the MMAT.

### Synthesis

#### Study quality

Overall, the quality appraisal found that few studies were empirically robust. Seven studies could not be appraised as they lacked an explicit study aim or research question, and therefore did not meet the Mixed Methods Appraisal Tool screening criteria for empirical studies^33, 34, 37, 40, 43, 46, 50^. Of the 14 that were appraised, most had moderate or low scores (three or lower, corresponding to meeting 60% of quality criteria or less), primarily due to either poor or poorly described study methodology. Mixed Methods Appraisal Tool scores for each study are presented in Table 2.

#### Telephone advice line characteristics (structures of care)

The 21 included studies featured 20 unique telephone advice lines, with one appearing in two studies (*Delivering Choice Programme’s* out-of-hours advice line^41, 42^) and one being reported post-implementation and re-evaluated after three years (*Palcall* telephone advice line^34, 37^). One further study^43^ reported two different telephone advice lines.

The main characteristics of the telephone advice lines in each study are provided in Table 3, with a summary of findings presented in Figure 2. Overall, 13 of the telephone advice lines were available for calls 24/7 and six were available during parts of the out-of-hours periods. One study did not state availability^36^. Fifteen of the telephone advice lines were accessible only for patients known to palliative care services (and their carers) or listed on palliative care registers, but information on how they were promoted or explained to patients and carers was uncommon. Information about the structure of the telephone advice lines was scant. Less than half of the studies described the location and coverage of the telephone advice line. The majority of services were not diagnosis-specific, with just one being restricted to cancer patients^51^. When reported, telephone advice lines were either financed by time-specific grants or funded internally by the service (Table 3).

**Figure 2.**
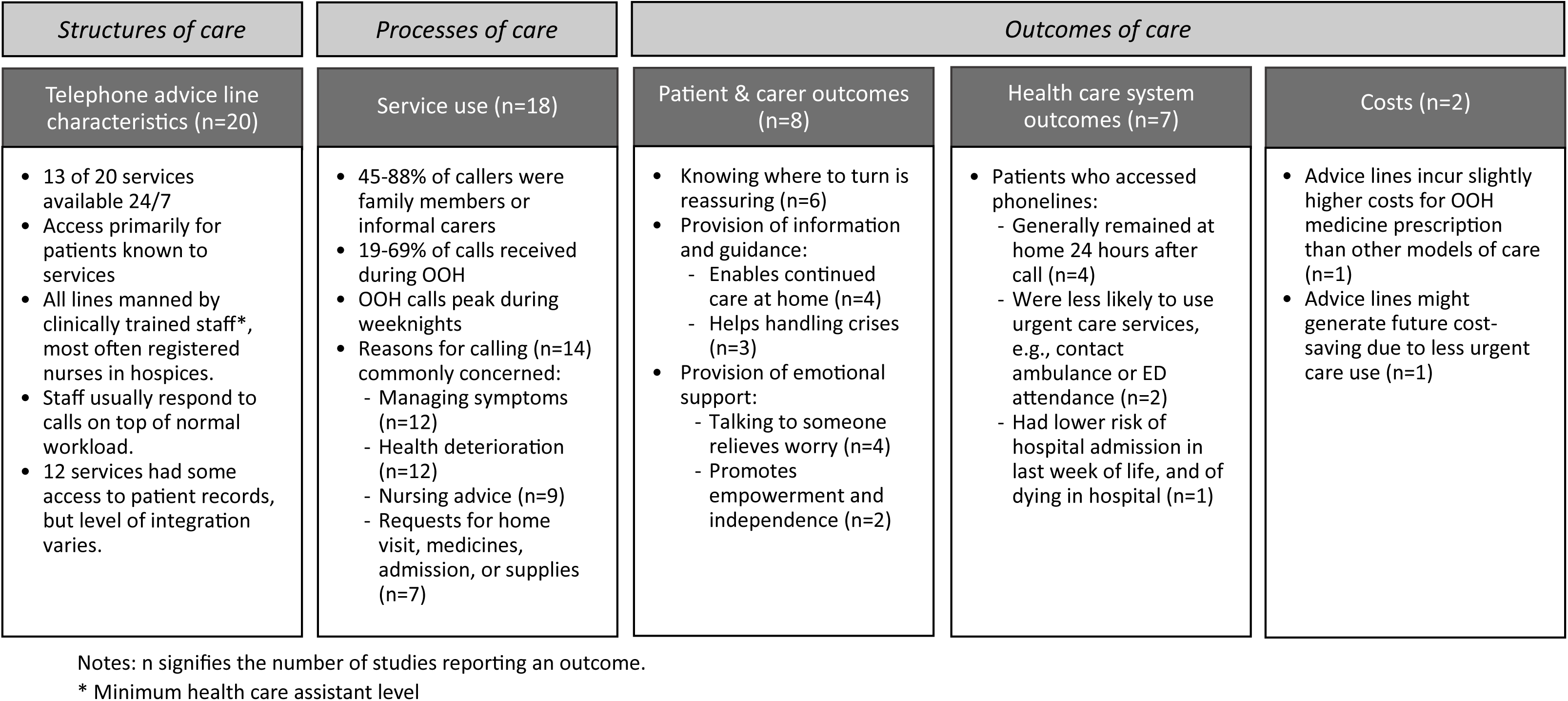
Summary of reported study findings applied to Donabedian’s^1^ framework of structures, processes, and outcomes of care.

**Table 3.**
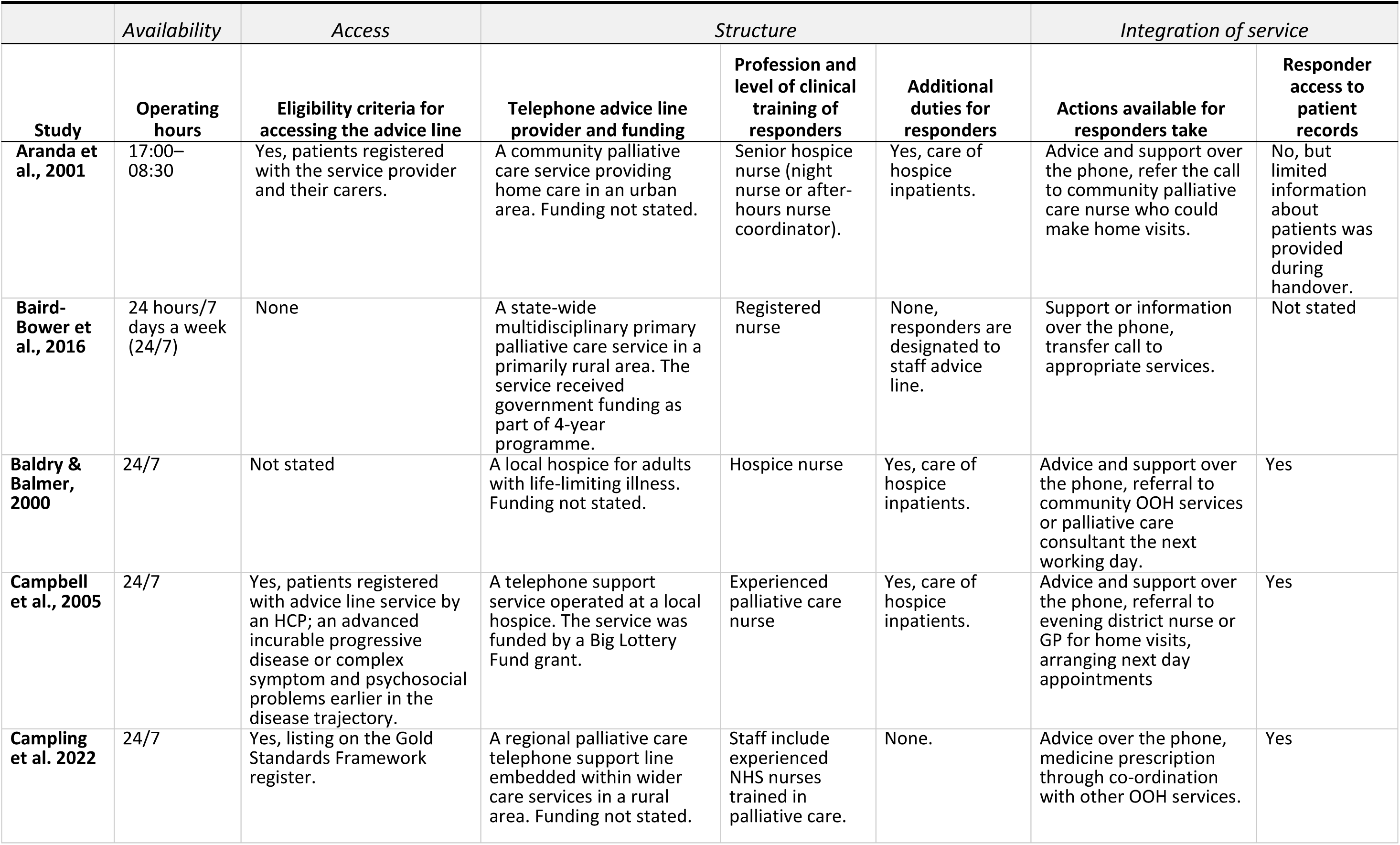
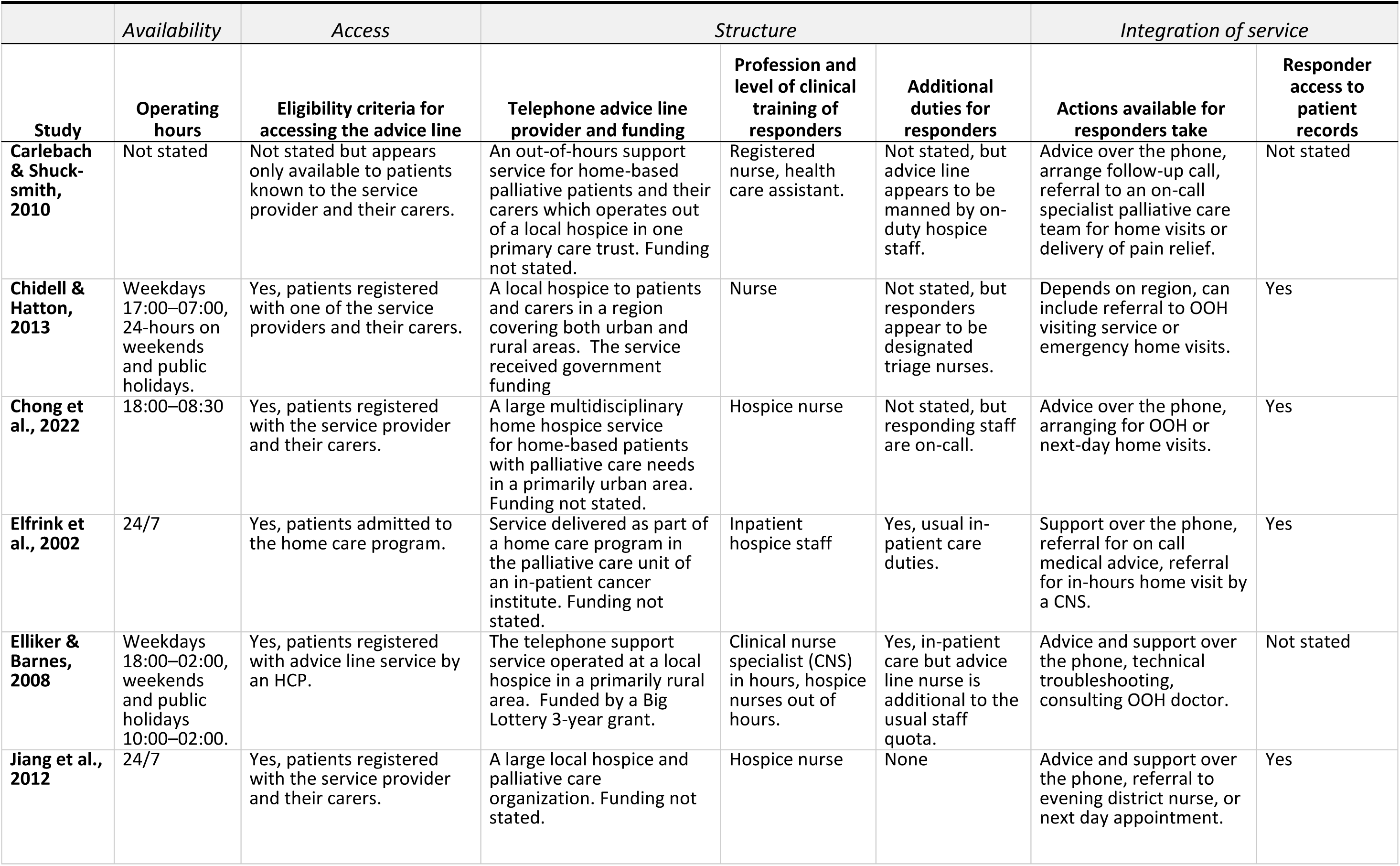
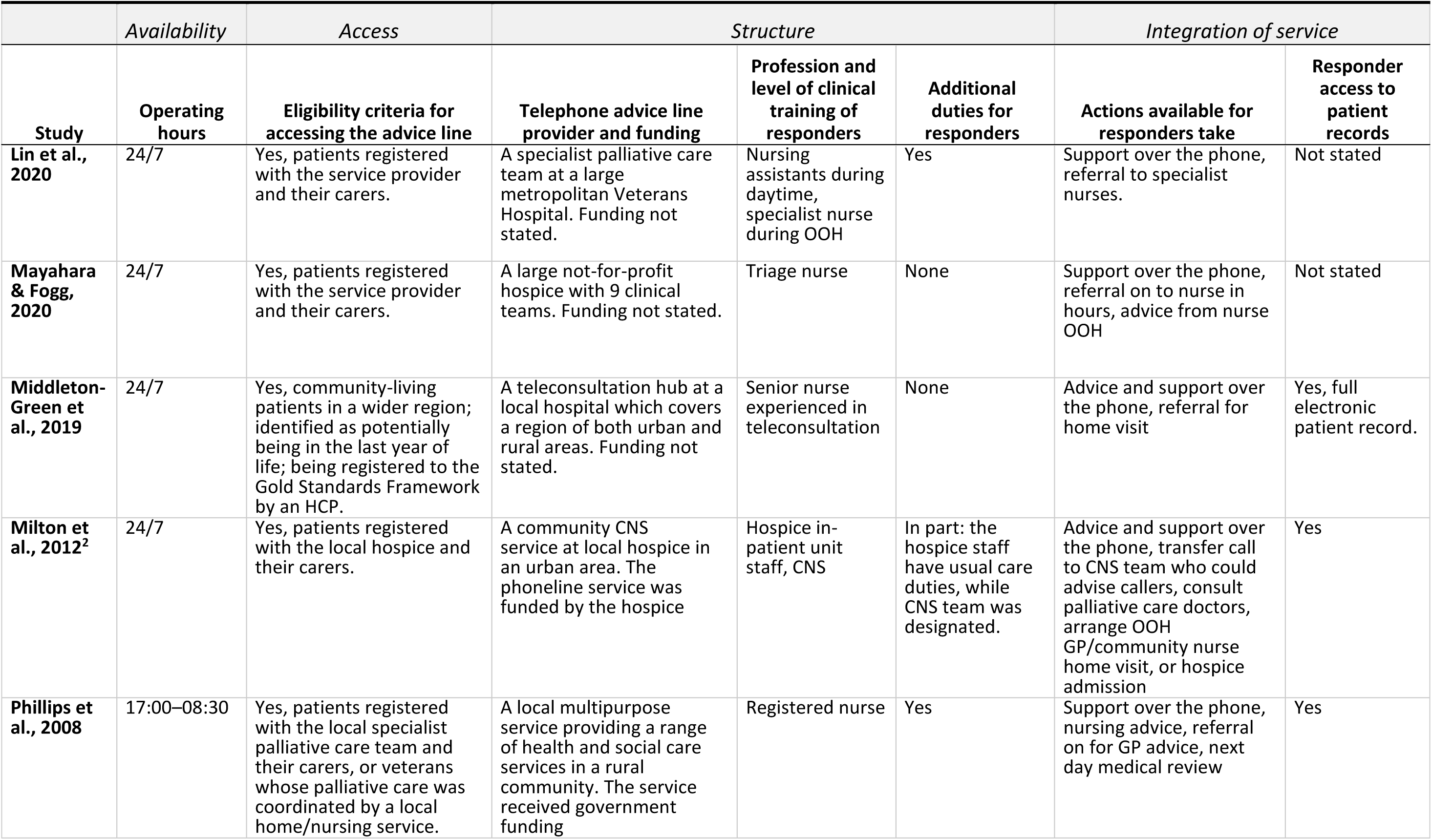
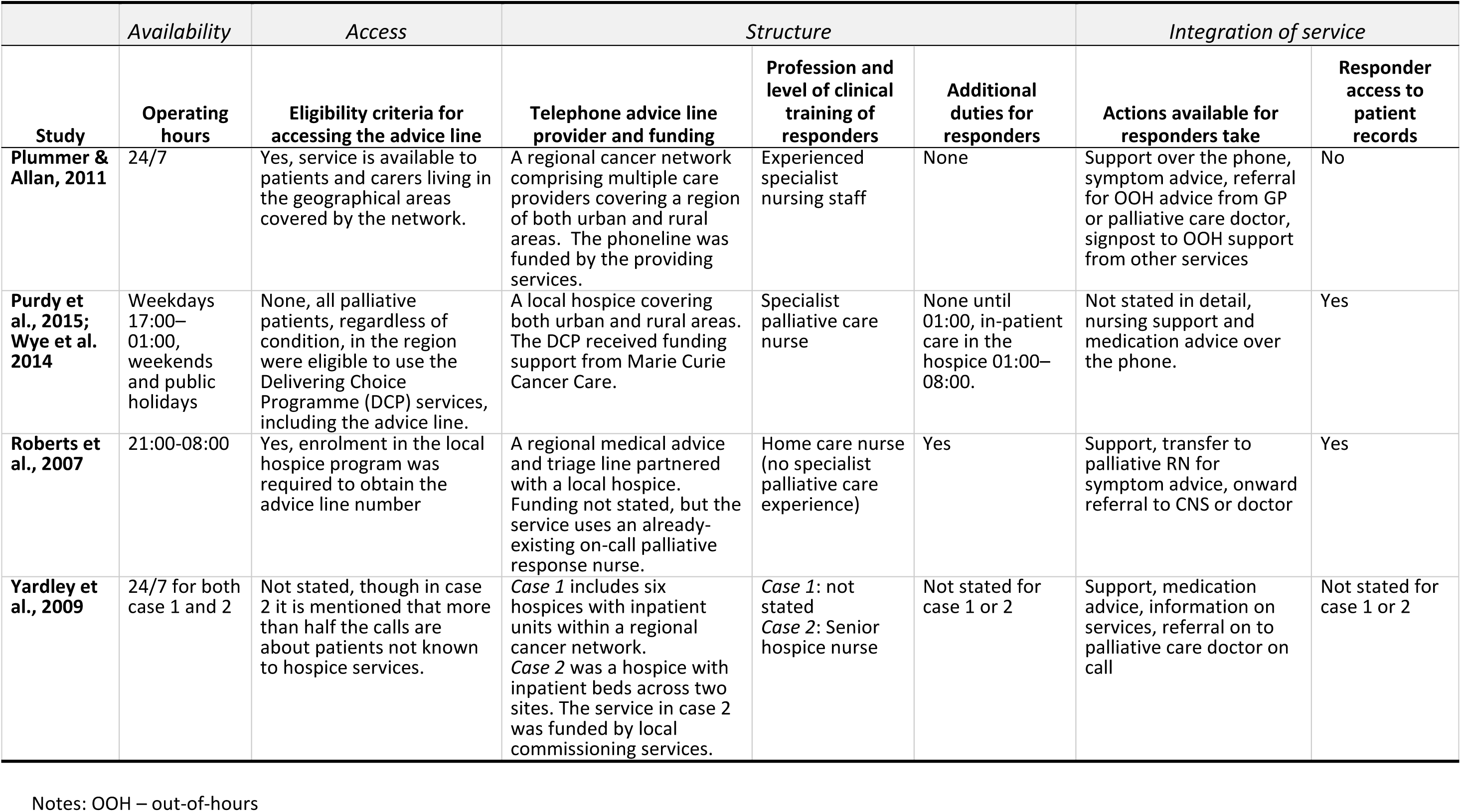
Characteristics of telephone advice line services reported in the included studies.

While the process of designing and implementing the telephone advice line service was described in several studies^34, 37, 39, 40, 43, 44, 47, 50, 51^, PPI or stakeholder consultation was rarely mentioned and only two studies reported involving patients and carers during development and feasibility testing of services ^34, 37^. Two further studies mentioned consulting key stakeholders during development but provided no detailed information on who or what this entailed^43, 47^.

#### Service use (processes of care)

Aspects of service use were reported in 18 of 21 studies, usually related to call volumes, timing of calls, identities of callers, reasons for calling, and information on how the call was handled by the responder or the organization (Figure 2).

In the seven studies that had 24/7 telephone advice lines and reported call data, between 19-69% of calls were received during out-of-hours periods^33, 35, 38, 43, 46, 52, 53^. Of these, most out-of-hours calls (35-78%) were received on weeknights before midnight, though the level of detail in reports differs^34, 36, 37, 39–42, 44, 45, 47–51^.

In the 12 studies that distinguished caller identities, callers were primarily identified as family members or informal carers (ranging 45-88% of calls) ^33, 34, 36, 37, 40, 45, 47, 48, 51–53^. Additional characteristics of out-of-hours callers, such as sociodemographic information, were presented in two studies. A study from Singapore by Chong et al. ^52^ reported gender (48.6% male), age (mean age 75.5 years), cancer diagnosis (80.9%), ethnicity (80.3% Chinese, 9.5% Malay, 5.2% Indian, 5.0% Other), and marital status (62.0% married, 26.9% widowed, 6.5% single, 4.6% divorced or separated). An American study by Mayahara et al. ^49^ reported ethnicity (patients: 73% White, 5% Black, 2% Asian, 20% Other; carers: 50% White, 25% Black, 25% Asian, 5% Other) and gender (60% female patients, 80% female carers), as well as diagnoses (most patients had ‘senile degeneration of the brain’ (n=212) and/or Alzheimer’s disease (n=116), heart failure (n=71), chronic obstructive pulmonary disease (n=58) or other advanced illnesses). Moreover, four studies reported basic demographics such as diagnosis, age, gender, or household composition for a wider patient sample registered with, or eligible to access, the service^38, 41, 44, 51^. Baird-Bower et al. ^45^ collected certain sociodemographic information of callers by linking call log data to patient records, but only reported callers’ location.

Reasons for calling were presented in 12 studies, with varying degrees of detail. The most reported call reason was managing symptoms and/or medication (*n*=10), with other reasons involving health deterioration, nursing advice, requests for new or additional medication or equipment, or reporting that the patient had died ^33, 34, 39, 40, 43, 44, 47, 48, 51, 52^. Since few studies provided information about follow-up periods, data about when in the care trajectory calls were made were scarce. Purdy et al. found that median time for accessing the telephone advice line was 10 days before death^41^ and Jiang et al. ^48^ reported that most calls occurred between the first days of home hospice admission and the last three days of life. Roberts et al. ^50^ and Middleton-Green et al. ^38^ presented qualitative data indicating that people call in the final days of life for advice and support when the patient is dying, as exemplified by this carer: “*I think I just asked the question ‘is this what I think it is?’ And I think all they said was ‘yes, probably’*” ^38^.

Fifteen studies gave some information about call handling. Actions available to responders differed between telephone advice lines and were variably described. Overall, the most reported actions taken by responders involved providing support and nursing advice appropriate to their level of clinical training, and transferring or signposting callers to other services. Responders in some telephone advice lines also contacted on-call teams for urgent home visits (n=5), arranged next-working-day follow-up at home or over the telephone (n=9), and, less commonly, helped with prescribing or collecting medication (n=4) (Table 3). Ten studies reported that responders successfully handled many calls directly through over-the-phone support^33, 34, 38, 40, 44, 46, 48, 50, 51, 53^. In cases when a responder could not adequately help a caller, calls were usually passed on to more senior healthcare professionals, commonly senior palliative care nurses, clinical nurse specialists, or GPs, who were either available on-call or in-house (n=11) ^33, 35, 38, 40, 42–44, 46, 48, 50, 51^. These professionals could generally provide more advanced medical advice, arrange home-visits, make referrals, and occasionally, prescribe or dispatch medicines. However, the proportion of calls passed on was rarely reported.

#### Effectiveness (outcomes of care)

Reported outcomes in the studies primarily related to patients’ and carers’ qualitative accounts of using a service and derived from open-ended survey questions or interviews rather than quantitative outcomes. A small number of studies used patient records to assess possible change in use of health care services and estimated costs or cost-savings in relation to expected benefits.

##### Patient and carer outcomes

No study provided quantitative data about effects on patient and carer outcomes. However, there was qualitative evidence of patient and carer reports of benefit. Patients and carers reported that the telephone advice lines provided general information, medical advice, and practical guidance about symptom management or care procedures^36, 38, 40, 42, 47^. This was exemplified by a carer in Phillips et al.’s study^47^ stating that *“You have the information but when you go to do it, it is really scary. It was so good to have someone just telling me I was doing the right thing”*. Furthermore, the same study found that accessing advice enabled carers to continue caring at home, and Carlebach and Shucksmith^36^ highlighted how this promoted independence for patients at home who would otherwise rely on family and friends.

A recurring finding was that the telephone advice lines provided valuable opportunity for patients and carers to ask about signs and symptoms and be informed about the disease trajectory or the dying process, so that callers can better understand whether these are ‘normal’ or expected^38, 42, 50^. This was exemplified by a carer quoted by Middleton-Green et al. ^38^:

> *”[It] helped, just that somebody could tell me what was, reassure me that what I was seeing and experiencing … it meant that I could just be in the moment with her. It gave me sort of control over my feelings, which ultimately meant that she was looked after better than she would have been”*

Receiving appropriate information was also reported to aid decision-making and help prevent unnecessary hospital admission and promote appropriate use of care services, as patients and carers noted that they might otherwise have contacted emergency care services^38, 42, 47^.

Some studies reported how telephone advice line staff would liaise with other services on patients’ behalf, which was advantageous when callers were requesting medication, care equipment, or home visits^35, 38^. Carlebach and Shucksmith^36^ highlighted callers sense of comfort when responders were familiar to the patient’s case. Similarly, access to patient records was described as valued by callers who did not have to re-explain the situation, as described by a carer quoted by Campling et al. ^35^: *“You don’t have to go through the rigmarole of explaining everything. They’ve got your records there; they can see clearly what’s going on…”*.

#### Patient and carer experiences

Patient and carer experiences of using the telephone advice lines were reported in eight studies using qualitative data^35, 36, 38–40, 42, 47, 50^. User experiences were overwhelmingly positive. Calls were generally triggered by a patient crisis or carer uncertainty. Both patients and carers emphasised feeling reassured by the knowledge that the service was available for them whenever they might need it, both in medical crises and when the caller was feeling uncertain or worried^36, 38, 40, 42, 47, 50^. In Milton et al.’ study^39^, feedback from a carer stated that “*There was comfort in knowing* [staff] *were on standby when you needed them;* [it] *made things easier, not being alone here*”. In demanding situations, only having one phone number to call to get direct access to experienced staff was perceived as helpful. Wye et al. ^42^, for example, presented a carer who noted that: “*I didn’t ever have to phone for ambulances or anything, all that was done and it wasn’t done through the GP or the district nurse… you could just phone one number*”.

##### Health care system outcomes

Seven studies reported outcomes relating to callers’ subsequent care service use but only two of these were comparative. These linked calls to routine data over a defined follow-up period. Both demonstrated that patients who accessed telephone advice lines were less likely to use emergency care services. Baird-Bower et al. ^45^ examined patient records and mortality data up to six months after service use and found that users were less likely to call for an ambulance (χ^2^=70.094, *p*<0.01) ^45^ or visit an emergency department (χ^2^=78.286, *p*<0.01) compared to people who never called. Purdy et al. ^41^ had a shorter follow-up period but found that emergency department attendance was lower among callers compared to a cohort without access to a telephone advice line service both 30 (OR 0.60, *p*=0.007) and 7 days before death (OR 0.34, *p*=0.003). They also showed that callers were less likely to be admitted to hospital in the last week of life (OR 0.44, *p*=0.005) and to die in hospital (OR 0.34, *p*<0.001) compared to patients who did not use the service^41^.

While there was little evidence for whether calls influence patients’ and carers’ subsequent care seeking behaviour, six studies demonstrated that, overall, only between 1.5-9.0% calls led to hospitalization or admission of patients^38, 39, 46, 48, 50, 52^. This suggests that hospitalisation is infrequent following out-of-hours calls, although the lack of data makes it hard to determine the effect of telephone advice lines on subsequent admission. Three retrospective cross-sectional studies reported that 73-95% of patients who contacted the telephone advice line remained at home 24 hours after the call^38, 40, 50^. There was, however, no further follow-up on actual place of death. In addition, Aranda et al. reported that 37% of all deaths during the study period occurred at home, but this was not temporally or causally linked to use of the telephone advice line^44^.

##### Cost effectiveness

There was very scant evidence regarding costs or cost effectiveness of telephone advice lines. Three studies presented rough estimations of costs associated with implementing telephone advice lines within a specific service, suggesting they were generally ‘resource-effective’ for providing palliative care during out-of-hours periods, but did not provide detailed cost breakdowns or include cost-savings related to potential benefits of the service^39, 47, 51^. Mayahara & Fogg^49^ argued that reducing avoidable out-of-hours home visits in response to, e.g., medication or equipment requests would save overall health system costs but did not provide a cost-analysis.

Two studies provided wider cost-benefit estimations, but both were based on estimated rather than observed impact. Baird-Bower et al. ^45^ estimated that use of the telephone advice line could lead to general cost savings associated with less use of avoidable emergency care for people who want to die at home, with projections of 28% lower emergency department attendance saving 121,647 Australian dollars and a 35% reduction in ambulance calls saving 112,700 Australian dollars.

Campling et al. ^35^, on the other hand, reported that for the purposes of improving out-of-hours access to medicines for patients at home, the telephone advice line model incurred slightly higher costs than other models of care (1-year cost of prescribing for an assumed English patient population with palliative care needs estimated to £60,075,302, compared to £49,483,750 for standard general practice prescriptions and £43,267,634 for clinical nurse specialist prescriptions).

## Discussion

Although telephone advice lines are recommended in palliative care policy and guidelines, and widely implemented at local levels, our study found very limited empirical evidence for their clinical and cost effectiveness. Most studies report process measures, e.g., service use and handling of calls, whereas the potential impact on patient and carer health, wellbeing, and caring capacity, or influence on subsequent service use and costs is rarely assessed in a rigorous manner.

Findings from the more methodologically robust studies suggest possible support for the clinical effectiveness of telephone advice lines, with use being associated with remaining at home following the call and less contact with emergency care services^38, 41, 45, 50^. However, most studies were observational, often of low methodological quality without control or comparator groups and can therefore not establish causality. Similarly, qualitative data indicate that the mechanism of action for reducing unnecessary or inappropriate emergency care use is the provision of care advice and emotional reassurance from trained staff, which enables patients and carers to better cope with care provision at home, if the patient prefers this. The possible influence on emotional wellbeing and caring capacity is a promising finding considering projected rises of home deaths and increasing work for informal carers when palliative care provision shifts towards being community-based^54, 55^, especially since anxiety has been shown to contribute strongly to urgent care-seeking behaviour^56^.

Telephone advice line models varied according to accessibility, structure of service provision, and integration with other care services. Limitations of the synthesised data, e.g., poor quality and inconsistent or insufficient reporting, prevented meaningful assessment of which telephone advice line models are most effective. Taken together, however, the included studies suggest that telephone advice lines might provide patients and carers at home with a viable alternative to emergency care when health deteriorates outside the normal working hours of care services, which is in line with other studies exploring reasons for contacting care services out-of-hours at the end of life^57, 58^.

Most of the evidence derived from local studies on telephone advice lines services that are accessible primarily to people known to the providing care services, which raises questions regarding equity of access, as the wider population of people with palliative care needs in the community remain under-served. While a few studies^41, 44, 52^ considered differences in the use of telephone advice lines according to diagnosis, no studies analysed patterns of use according to socioeconomic position, ethnicity, or those with limited language proficiency. Language barriers in particular constitutes a key challenge associated with worse quality of care^59^ and higher rates of emergency care use at the end of life^60^.

Another gap in the existing literature relates to integration with other services, e.g., the extent to which patient records were accessible to staff responding to calls and shared with other health care professionals. Inadequate integration with the wider health care system has previously been identified as contributing to inequity in access to community-based palliative care and may limit benefits for patients and carers^61^. While it was not possible to assess whether access to patient records influences the effectiveness of telephone advice lines, the included studies suggest that such access to records allows responding staff to better understand the patient’s circumstances and provide appropriate advice.

Several of the included articles were based on internal audits rather than empirical research studies, which limits the generalisability of the evidence. In addition, most articles only encompassed service users, who might not be representative of the larger population of home-based adults with palliative care needs. Another potential source of bias is that authors in eight of the included articles were evaluating telephone advice lines services in which they had clinical or managerial roles^62^. The additional finding that patient and public involvement during stages of planning, designing, and evaluating telephone advice lines processes was uncommon, or at least rarely reported, further emphasises that the current literature relies heavily on the professional perspective. Inadequate use and reporting of patient and public involvement makes it difficult to gauge whether existing telephone advice lines are meeting the needs and preferences of the patients and carers they intend to serve. Involving patients and carers in co-design and evaluation is imperative to ensure relevance and patient-centred care delivery and will help identify hindering and facilitating factors for use^13, 63^. Use of guidelines, such as Guidance for Reporting Involvement of Patients and the Public^64^ in future studies is strongly recommended.

### What this review adds

Our findings have important implications for policy and research. The insufficient evidence base for telephone advice lines that we identify here stems largely from a lack of comprehensive and robust evaluations. The predominance of descriptive results (focusing on who calls, when and why they call, and how calls are handled by responding staff) reflects findings from previous reviews of telehealth^65^ and digital technologies used in palliative care practice without a strong evidence base^66, 67^.

To determine the effectiveness of telephone advice lines on an individual and population level in relation to costs, prospective studies with longer follow-up periods and linked to routine data identifying service use^68^ are necessary. Adhering to such standards would promote more rigorous and comparable research and help build an evidence base for effectiveness. Recommendations for key components of structure and use of telephone advice lines to record and report in future evaluations are presented in Supplementary file 2.

### Strengths and limitations

A major strength of this review is its relevance and rapidity. Finding specific and evidence-based ways to improve community-based palliative and end of life care is important if the growing demand for community-based care is to be met. Telephone advice lines are currently garnering much interest in policy and practice and it is important to provide a synthesised evidence-base to inform future development and implementation of these services. The decision to include all relevant articles identified in our search, regardless of their quality appraisal also strengthens the study, as we could demonstrate the poor scientific standard of the existing literature, with a third of the identified studies not judged as reaching the criteria of empirical research. By only including peer-reviewed articles, however, we might have missed relevant evaluations of additional telephone advice lines in published but not peer-reviewed reports, which is a limitation. The rapid review methodology resulted in searching a limited number of databases and supplementary searching of grey literature or forward and backward citation searching was not utilised. However, the search strategies were carefully designed and tested against known relevant studies by an experience information specialist^69^. Though most of the abstract and title screening was conducted by a single reviewer, which might increase the risk of missing relevant studies, single screening has been found to be equally sensitive for providing a sufficient evidence base in studies with focused clinical review questions as dual screening^70^.

Despite the rapid approach of this review, we incorporated discussions with PPI representatives with lived experience of end-of-life care at several stages of the study process, using both one-time and continuous approaches^32^, to guide data extraction, synthesis, and presentation of results. This enhances the relevance of the study for patients and carers.

## Conclusion

Despite being recommended in policy and increasingly used in community-based palliative care services, evidence for the clinical and cost effectiveness of telephone advice lines is limited. Existing evidence primarily constitutes descriptive reports of service use and service structures, and qualitative accounts of carers’ and patients’ experiences of using telephone advice lines. Overall, our findings suggest that telephone advice lines provide guidance and reassurance that can support care at home and reduce avoidable emergency care use in the last months of life. The variability in reporting, and poor methodological quality, across studies hindered meaningful synthesis evidence regarding patient/carer and health care system outcomes. Development of new telephone advice line services ought to include patients and carers to ensure that needs and preferences of intended users are considered, and our provided recommendations for more rigorous evaluations should be followed to strengthen the evidence base.

## Supporting information

Supplement file 1. Search strategies

Supplement file 2. Recommendations

## Supplement files

**S1. Search strategies**

Separate document.

**S2. Recommendations for future evaluations.**

Separate document.

## Declarations

### Authorship

T.J. was responsible for the design of the study, with critical input from all authors. S.G., T.J., and S.P. developed the search strategy and S.G conducted the searches. T.J., T.C., R.C., M.B. and A.B. were screened articles for inclusion, extracted data, and performed quality appraisal. All authors contributed to analysis and interpretation of data. T.J. was responsible for drafting and revising the manuscript. All authors critically revised the manuscript for intellectual content, and read and approved the final manuscript.

### Ethics and consent

Ethical approval was not required for this review as there was no direct patient contact or access to individual participant data.

### Data sharing

This review is registered on the PROSPERO database (CRD42023400370, available at crd.york.ac.uk/prospero/display_record.php?RecordID=400370) and the full search strategy is provided in the Supplementary material. Inclusion/exclusion criteria and quality appraisal results are available in the article. Additional data will be made available upon request.

## Footnotes

1 Donabedian. The quality of care: how can it be assessed? *JAMA*. 1988; 260: 1743-8.

