## Supplement file 1. Search strategies for "A rapid systematic review of the effectiveness of out-of-hours palliative care telephone advice lines for people living at home and their carers"

### S1. Search strategies

All searches performed on 09 February 2023

#### Medline All via OVID

(Ovid MEDLINE(R) ALL 1946 to February 08, 2023)

1.	exp advance care planning/

2.	exp attitude to death/

3.	exp bereavement/

4.	death/

5.	hospices/ or "Hospice and Palliative Care Nursing"/

6.	life support care/

7.	palliative care/ or Palliative Medicine/

8.	exp terminal care/ or respite care/

9.	terminally ill/

10.	palliat$.af.

11.	hospice$.af.

12.	(terminal care or respite care).af.

13.	or/1-12

14.	journal of palliative care.jn.

15.	journal of palliative medicine.jn.

16.	hospice journal physical psychosocial & pastoral care of the dying.jn.

17.	supportive care in cancer.jn.

18.	palliative medicine.jn.

19.	palliative & supportive care.jn.

20.	journal of supportive oncology.jn.

21.	journal of social work in end of life & palliative care.jn.

22.	journal of pain & symptom management.jn.

23.	journal of pain & palliative care pharmacotherapy.jn.

24.	international journal of palliative nursing.jn.

25.	death studies.jn.

26.	death education.jn.

27.	american journal of hospice care.jn.

28.	american journal of hospice & palliative medicine.jn.

29.	omega journal of death & dying.jn.

30.	or/14-29

31.	13 or 30

32.	bereave*.mp.

33.	attitude to death.mp.

34.	end of life.af.

35.	Advance* Care.af.

36.	((advanced or terminal*) adj (ill* or disease)).ti,ab,kw.

37.	supportive care.ti,ab,kw.

38.	dying.ti,ab,kw.

39.	"last year of life".ti,ab,kw.

40.	(limited life adj (expectanc* or span*)).ti,ab,kw. or life-limiting.mp.

41.	or/32-40

42.	31 or 41 [palliative care concept]

43.	exp After-Hours Care/

44.	Night Care/

45.	(after hour* or ((outside or out or after or off) adj2 (normal or working or office) adj2 (time or hour*))).ti,ab,kw.

46.	after office hour*.ti,ab,kw.

47.	(out of hours or (OOH or OOHs)).ti,ab,kw.

48.	out of office hours.ti,ab,kw.

49.	(off adj hour*).ti,ab,kw.

50.	((weekend* or evening* or holiday* or night*) adj (hour* or care*)).ti,ab,kw.

51.	(((24 hour* or 24H or around-the-clock or around the clock) adj2 (care or access* or service* or support*)) or "24 hours per day").mp.

52.	or/43-51 [OOh concept]

53.	(hotline* or advice line* or telephone support or telephone helpdesk* or helpline*).mp. or Hotlines/ [hotlines]

54.	exp Telephone/

55.	(phone* or telephone*).mp.

56.	Telemedicine/

57.	(telehealth* or telemedicine or telehospice* or tele-health or tele-medicine or videophone* or video phone*).mp.

58.	or/54-57 [broad telephone/telemedicine]

59.	52 and 58 [OOH AND broad telephone/telehealth]

60.	53 or 59 [hotlines OR (OOH telephone/telehealth)]

61.	("17555161" or "16471046" or "17169964" or "12802979" or "19568212" or "11510411" or "22790013" or "22068119" or "24838731" or "20871499" or "26850118" or "23123986" or "18411012" or "3966617" or "19702614" or "12148971" or "11309907" or "16438809" or "21378067" or "15200518" or "24950521" or "27349847" or "22773920" or "17165090" or "15188918" or "12699575" or "15022976" or "18928135" or "18788963").ui. [29 relevant studies]

62.	42 and 60 [palliative care AND (hotlines OR OOH telephone/telehealth)]

63.	61 or 62

#### Embase via OVID

(Embase 1974 to 2023 February 08)

1.	advance care planning/

2.	attitude to death/

3.	bereavement/

4.	death/

5.	hospice/

6.	exp palliative therapy/

7.	respite care/

8.	terminal care/ or hospice care/

9.	exp terminally ill patient/

10.	palliat$.af.

11.	hospice$.af.

12.	(terminal care or respite care).af.

13.	supportive care.ti,ab,kw.

14.	bereave$.mp.

15.	attitude to death.mp.

16.	end of life.af.

17.	((advanced or terminal* or critical*) adj (ill* or disease)).ti,ab,kw.

18.	Advance* Care.af.

19.	(limited life adj (expectanc* or span*)).ti,ab,kw. or life-limiting.mp.

20.	"last year of life".ti,ab,kw.

21.	dying.ti,ab,kw.

22.	or/1-21 [palliative concept]

23.	out-of-hours care/

24.	night care/

25.	(after hour* or ((outside or out or after or off) adj2 (normal or working or office) adj2 (time or hour*))).ti,ab,kw.

26.	after office hour*.ti,ab,kw.

27.	(out of hours or (OOH or OOHs)).ti,ab,kw.

28.	out of office hours.ti,ab,kw.

29.	(off adj hour*).ti,ab,kw.

30.	((weekend* or evening* or holiday* or night*) adj (hour* or care*)).ti,ab,kw.

31.	(((24 hour* or 24H or around-the-clock or around the clock) adj2 (care or access* or service* or support*)) or "24 hours per day").mp.

32.	or/23-31 [OOH concept]

33.	hotline/

34.	(hotline* or advice line* or telephone support or telephone helpdesk* or helpline*).mp.

35.	33 or 34 [hotlines]

36.	telephone/

37.	(phone* or telephone*).mp.

38.	telemedicine/ or telehealth/

39.	(telehealth* or telemedicine or telehospice* or tele-health or tele-medicine).mp.

40.	(videophone* or video phone*).ti,ab,kw.

41.	teleconsultation/

42.	or/36-41 [broad telephone/telemedicine]

43.	32 and 42 [OOH and broad telephone/telemedicine]

44.	35 or 43 [hotlines or OOH+telephone]

45.	22 and 44 [palliative care AND hotlines/OOH telephone]

46.	limit 45 to conference abstract status

47.	45 not 46

48.	limit 47 to (books or editorial)

49.	47 not 48 [remove books, editorials]

#### CINAHL via Ebsco

| **#** | **Query** | **Results** |
| --- | --- | --- |
| S1 | ( (MH "Terminal Care+") or (MH "Palliative Care") or (MH "Attitude to Death") or (MH "Advance Care Planning") or (MH "Respite Care") or (MH "Hospices") or (MH "Life Support Care") ) OR TI ( bereave* or hospice* or "end of life" of "terminally ill" or palliat* ) OR AB ( bereave* or hospice* or "end of life" of "terminally ill" or palliat* ) | 110,127 |
| S2 | TI life-limiting OR AB life-limiting | 1,929 |
| S3 | TI ( ((advanced or terminal* or critical*) n1 (ill* or disease)) ) OR AB ( ((advanced or terminal* or critical*) n1 (ill* or disease)) ) | 46,351 |
| S4 | TI ( (limited life N1 (expectanc* or span*)) ) OR AB ( (limited life N1 (expectanc* or span*)) ) | 503 |
| S5 | S1 OR S2 OR S3 OR S4 | 150,792 |
| S6 | TI out of hours or OOH or OOHs OR AB out of hours or OOH or OOHs | 2,779 |
| S7 | (MH "Night Care") | 488 |
| S8 | TI ( ((outside or out or after or off) N2 (normal or working or office) N2 (time or hour*))) ) OR AB ( ((outside or out or after or off) N2 (normal or working or office) N2 (time or hour*))) ) | 558 |
| S9 | TI "after hour*" OR AB "after hour*" | 970 |
| S10 | TI out of hour* OR AB out of hour* | 2,750 |
| S11 | TI "off hour*" OR AB "off hour*" OR TI ( ((weekend* or evening* or holiday* or night*) N1 (hour* or care*)) ) OR AB ( ((weekend* or evening* or holiday* or night*) N1 (hour* or care*)) ) | 1,616 |
| S12 | TI ( ((24 hour* or 24H or around-the-clock or around the clock) N2 (care* or service* or support*)) ) OR AB ( ((24 hour* or 24H or around-the-clock or around the clock) N2 (care* or service* or support*)) ) | 563 |
| S13 | S6 OR S7 OR S8 OR S9 OR S10 OR S11 OR S12 | 6,550 |
| S14 | (MH "Telephone Information Services") | 3,490 |
| S15 | TI ( hotline* or advice line* or telephone support or telephone helpdesk* or helpline* ) OR AB ( hotline* or advice line* or telephone support or telephone helpdesk* or helpline* ) | 3,777 |
| S16 | S14 OR S15 | 6,377 |
| S17 | (MH "Telehealth+") OR (MH "Telephone") OR (MH "Videoconferencing") | 53,206 |
| S18 | TI ( phone* or telephone* or telehealth* or telemedicine or telehospice* or tele-health or tele-medicine or videophone* or video phone* ) OR AB ( phone* or telephone* or telehealth* or telemedicine or telehospice* or tele-health or tele-medicine or videophone* or video phone* ) | 69,315 |
| S19 | S17 OR S18 | 99,763 |
| S20 | S13 AND S19 | 602 |
| S21 | S16 OR S20 | 6,880 |
| S22 | S5 AND S21 | 175 |
