## Supplement file 2. Recommendations for "A rapid systematic review of the effectiveness of out-of-hours palliative care telephone advice lines for people living at home and their carers"

### S2. Recommendations for future evaluations

Recommendations for information to record and report, in addition to any outcome measures, to describe models of telephone advice line services in future evaluations.

#### Development of service

- What is the intended purpose of the phoneline?
- Have patients and carers been involved in shaping and refining the service?

#### Access to service

- Who is eligible to call and how is information about the phoneline advertised?

#### Structure of service

- How is the phoneline funded and which service is responsible for/manages it?
- Who responds to calls, what is their profession and level of clinical training?
- Have responding staff received training in telephone triaging, communication skills, etc.?
- Are responding staff dedicated to taking calls or responding to calls on top of their usual workload?
- Do responders have access to medical records and can they update these?

#### Availability of service

- What are the operating hours of the service?

#### Service use

- Does the call concern a person who is known to the service or not (i.e., registered with the providing service or listed on a palliative care register)?
- Is the caller a:
  i. person with palliative and end-of-life care needs
  ii. family member or informal carer
  iii. a professional carer (incl. home care)
- The timing of the call, at least distinguishing calls received:
  - in-hours (morning/afternoon)
  - out-of-hours (weekday evenings and nights, weekends, and public holidays)
- The main reason for the call
- The outcome of the call:
  - Advice and support over the phone which solved the query/issue
  - Transferring caller directly to another health care professional
  - Dispatching on-call staff (in-house or through other services) for a home visit/medication or equipment dispense
  - Contacting emergency services
  - Arranging a next-working day follow-up

Signposting to other services
